# Projected Cancer Burden, Challenges, and Barriers to Cancer Prevention and Control Activities in the State of Telangana

**DOI:** 10.1101/2022.11.16.22282378

**Authors:** Hemant Mahajan, Reddy M Neha, NG Marina Devi, Usha Rani Poli, M Jayaram, Shailaja Tetali, GVS Murthy

**Affiliations:** Indian Institute of Public Health, Hyderabad, India; Department of Clinical Research, London School of Hygiene & Tropical Medicine, London, UK; MNJIO & RCC, Hyderabad, India; KIMS-ICON Hospital, Visakhapatnam, India; Apollo Hospitals, Chennai, India; Lilavathi Hospital, Mumbai; Medical Oncologist, Krishna Institute of Medical Sciences, Hyderabad, India; National Institute of Mental Health & Neurosciences, Bangalore, India; Apollo Cancer Hospital, Hyderabad, India

**Keywords:** Access, Cancer, Healthcare system, India, Magnitude, Telangana

## Abstract

**Background and Aim:** The Telangana cancer care program is a proactive, comprehensive initiative encompassing infrastructure development, human resource skilling and ensuring financial protection to those below poverty line. The broad aim of this exercise was to identify modalities to augment the Telangana State Cancer control Plan to implement a sustainable comprehensive cancer care model for Telangana.

**Methods:** We conducted in-depth interviews of stakeholders (17 patients and 25 providers) to identify barriers and challenges to access existing cancer care system; calculated the estimated magnitude of cancer and commensurate workload (in terms of visits to tertiary cancer care system for chemotherapy, radiotherapy, and surgery and human and equipment requirement) for the next 15 years (from 2021 to 2036). Using the anecdotal evidence and information from stakeholders’ interviews, we developed patient-journey funnels for cancer patients, which helped us to appreciate at what levels of care leakages occur.

**Results:** We estimated a 28% increase in the number of new cancer cases per year and the resultant workload: number of visits, chemotherapy, radiotherapy, surgeries, specialized human resources and equipment, from 2021 to 2036. Stakeholders mentioned delayed access to healthcare system as the main reason for the poor prognosis of patients. The common reasons cited for delayed access were: poor cancer-literacy including prevailing myths and misconception, financial barriers, and rural residence. Patient journey funnel for cancer care revealed major leakage from screened positive to diagnosis confirmation step. Patient leakage varies from ∼70% to 90% from screened positive till treatment completion.

**Conclusion:** Govt. of Telangana has initiated several measures to strengthen the healthcare system and to promote the uptake of cancer care services to manage the rising burden of cancer and resultant increasing workload. However, there is ample scope for further improvement (such as improved healthcare access, reduced patient leakage, commensurate human skills and infrastructure development etc.) to deliver comprehensive cancer care services in the state.

**Highlights:**

- Cancer cases are rising in Telangana, which is experiencing high epidemiological transition
- The healthcare infrastructure for cancer care is predominantly urban-centric
- Overall, 28% increase in new cancer cases and resultant workload per year from 2021 to 2036 was estimated
- Delayed access to healthcare system due to barriers at community and health system level
- Major patient leakage across cancer care continuum, which ranges from 70 to 90%

## Introduction

The burden of cancer is rising across all states in India at variable pace. This heterogeneity of cancer burden and/or health loss (across states) is due to differences in population genetics, social development, lifestyle, and environment. Owing to epidemiological transition [1] (the ratio of all-age disability-adjusted life years (DALYs) due to communicable diseases versus those due to non-communicable diseases (NCDs) and injuries together), the greatest increase in crude cancer incidence rates has been observed in states with high epidemiological transitions (an increase in the proportion of disease burden attributable to NCDs) while the mortality-to-incidence ratio (defined as number of deaths divided by number of newly diagnosed cancer cases in a given year) is greater in the other regions [2]. It has been reported that the age-adjusted cancer DALYs vary by 2.6 times across the states in India, which emphasize the need for contextualized state-specific interventions to handle the spiraling burden [2].

Telangana (TS), one of the southern states in India, has experienced a recent surge in number of cancer cases with crude-cancer incidence rate of 72.6 (69.4, 77.3) and age-adjusted incidence rate of 88.7 (85.1, 94.0) per 100,000 in 2016 [2]. The state-specific crude cancer mortality-to-incidence ratio was estimated to be 0.70 for females and 0.81 for males, which indicates poorer survival especially among males [2]. Of all the five neighboring states, cancer-related mortality-to-incidence ratio among females as well as males is only lower than Andhra Pradesh and Odisha. Among all population-based cancer registries in India, Hyderabad district (the capital of Telangana) recorded the highest incidence rate for breast cancer (leading cause of cancer mortality for women in Telangana) - 48/100,000 [3]. Moreover, two thirds of the cases of triple negative breast cancer (aggressive form of breast cancer with poor prognosis) were reported among participants aged less than 50 years [4]. Among men, the cancers with highest prevalence in the Hyderabad district were mouth, lung, and the tongue cancers [3].

The problem of the high burden of cancer in Telangana is further complicated by inadequate healthcare infrastructure (such as inadequate human resources, equipment etc.) due to lack of state-run health institutes dedicated for cancer management especially at primary and secondary level [5, 6]. This highlights a need to strengthen the healthcare system in TS to facilitate the comprehensive cancer care, especially in rural areas (which is unequally experiencing the cancer risk factors and preventable morbidity, mortality, and sufferings from cancers compared to their urban counterparts) [7-9]. Therefore, we conducted an assessment of the current control initiatives to identify potential public health value additions to the existing cancer care initiative of the state and strengthen it further. The broad aim of the initiative was to complement the excellent cancer care initiatives of the government of Telangana by building patient-centric pathways for comprehensive cancer care alongside developing academic and research capabilities in the state. The specific objectives were (a) To identify measures to-strengthen Telangana State comprehensive cancer care plan contextualized to local priorities and learnings from global best practices/models; (b) To co-create tangible interventions for implementation at the state level; and (c) To develop a robust advocacy plan and engage with policy makers.

In this paper we are presenting (a) cancer-specific burden and the commensurate workload for the state of Telangana for next 15 years (from 2021 to 2036); and (b) The findings from the stakeholder interviews at different levels of health care system to explore their perspectives on barriers and facilitators for comprehensive cancer care services in the state.

## Materials and methods

The work on the project was done between February and September 2021. The project was approved by the Institutional Ethics Committee of the Indian Institute of Public Health – Hyderabad. Data was extracted from published and grey literature to assess the magnitude of cancers, the determinants and barriers to access services, skills and workload, technology innovations in the continuum of cancer care, patient journey, palliative care services and successful cancer care interventions in other states in India and other parts of the world.

### Assessment of magnitude of cancer

For estimating the cancer-specific burden in Telangana, five-year prevalence of cancer in India as reported by Global Cancer Observatory, 2020 was considered. Due to the absence of data on cancer-prevalence in Telangana in 2021, the available proportions for India were applied and prevalence for Telangana was computed. The estimated population of Telangana for 2021 to 2036 was retrieved from the ‘ Report of the technical group on population projections, 2019’ [10].

Crude incidence rates of all cancers as estimated by the Global Burden of Disease study group for states of India in 1990 and 2016 were used to project annual incidence from 2021 to 2036 in Telangana. With the assumption that annual percentage change (APC) in crude incidence rate would remain the same for all states in higher-middle epidemiological transition level, the APC for all cancers in Telangana was considered to be 0.013 (0.009, 0.016) [2].

Assuming the overall survival rate to be 90%, cases surviving at the end of each year were estimated which were assumed to be the baseline for the subsequent year. Similarly, crude incidence rates for breast, cervical, oral and lung cancers over the years were projected. For common cancers (breast, cervical and oral) and lung cancer, the APC in age-standardized incidence rate was used as information on APC in crude incidence rate could not be traced. Survival rates for India were applied to determine magnitude of burden from each of these cancers in Telangana [11].

### Assessment of magnitude of cancer Workload

For understanding the workload for managing different cancers, the magnitude of cancers alone will underestimate the workload, as cancer patients have to make a number of visits for the surviving period with cancer for treatment and follow up. The probable number of visits for common cancers (oral, breast, cervical, and lung) was computed using expert opinion. Workload at the tertiary cancer centers was estimated based on the site and stage of cancer. Data on relative proportion of cases diagnosed in early and advanced stages was extracted from the Report on the National Cancer Registry Programme-2020 [3] which presented information on stages at diagnoses for specific cancers from hospital-based cancer registries across India. Average number of visits required during each phase (pre- and post-diagnosis) were determined based on interviews with key stakeholders (oncologists) of both public and private hospitals.

- Work-load was estimated using the formula
  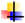Work-load = New case **×** visits per case during active treatment
- Demand for human resources (cancer specialists) and equipment for the respective years was estimated using the formula [12]:
  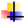No. = Estimated New Cases in the respective year **×** Norm per 1000 new patients
- Based on estimated new cases in the respective years, cancer cases requiring chemotherapy, radiotherapy and surgery were calculated for three assumed scenarios: 55%, 60%, 65% of new cases requiring chemotherapy and radiotherapy; and 45%, 40%, and 35% of new cases requiring surgery.
  - For each of the scenarios, total cases requiring chemotherapy was calculated using the formula:
    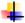Total cases requiring chemotherapy = [Estimated new cases in a given year **×** Percentage requiring chemotherapy]
  - To calculate chemotherapy cycles per day, assuming six cycles are required per patient, the formula used was as follows:
    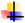Chemotherapy cycles per day = [New case requiring chemotherapy in a given year **×** 6]/365
  - Similarly, for estimating linear accelerators for radiotherapy sessions, we assumed 22 sessions per patient and 15,000 sessions per machine. The following formula was applied:
    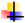Linear accelerators for radiotherapy sessions = [Estimated new cases requiring radiotherapy in a given year in a given scenario **×** 22]/15000
  - Onco-surgeries that are likely to be performed a month were estimated by applying the formula
    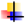Onco-surgeries that are likely to be performed a month = [Estimated new cancer cases requiring surgery annually in each scenario]/12

### Interviews with stakeholders

In-depth interviews were conducted with various stakeholders (n= 25) serving at different levels of healthcare system in different capacities along with 17 patients with different sites of neoplasms and different stages to explore their perspectives on barriers and facilitators of comprehensive cancer care services in the state. A non-probabilistic purposive sampling technique was applied to identify oncologists, medical officers at primary health centers (PHC), community health centers (CHC) and district hospital (DH), staff-nurses, auxiliary nurse and midwife (ANMs), mid-level health provider (MLHP), palliative care physicians, district programme officer of National Program for the prevention and control of cancer, diabetes, cardiovascular diseases and stroke (NPCDCS), and administrative secretary of a non-governmental organization. An informed verbal consent was obtained prior to every interview. The interview-probes were developed by the investigators in consultation with experts. Similarly, patients diagnosed and either seeking or receiving treatment for different cancers were identified through convenient sampling (n= 17) at three tertiary cancer care centers in Hyderabad and interviewed after informed verbal consent.

We developed patient journey funnels for cancer, which had helped us to appreciate at what levels of care leakages occur. To develop this, a number of assumptions were made (most of which were anecdotal and not evidence-based). We developed patient journey funnels for three common cancers together (oral, breast, and cervical) as well as separately for breast and cervical cancers. We calculated patient leakages at various points and pathways across cancer care continuum such as (i) Screened positive to diagnosis confirmation; (ii) Diagnostic confirmation to treatment initiation and treatment completion; (iii) Screened positive till treatment completion; and (iv) Diagnosis till treatment completion.

For the quantitative data such as workload, magnitude etc., we calculated descriptive statistics (as appropriate) and 95% using Binomial Exact using STATA version 14.2 (Stata Corp, College Station, TX, USA). For the analysis of qualitative data, open coding was done and themes were generated. The frequencies of codes were used for further analyses.

## Results

Table 1 describes the magnitude and corresponding workload for oral, breast, cervical, and lung cancers from 2021 to 2036. Approximately, 27% increase was anticipated in the crude incidence of breast cancer between 2021 and 2036. A 22% decrease was projected in the crude incidence of cancer of the cervix between 2021 and 2036. No change is anticipated in the crude incidence rate of oral cancers. Regarding lung cancer no significant changes in crude incidence rates per 100,000 or magnitude is anticipated, despite demographic changes till 2036. It was estimated that for early-stage breast cancers, the workload will increase by nearly 33% between 2021 and 2036 (Table 1). Similarly, ∼18% decrease in workload was anticipated between 2021 and 2036 for cervical cancer. However, significant differences were not anticipated for either Oral cavity cancers or Lung cancer.

**Table 1.** Estimated work load for the breast, cervical, oral, and lung cancer for the next 15 years

| Year | CIR | New cases | Early stage | Late stage | Workload | Survival after 5<sup>th</sup> year |
| --- | --- | --- | --- | --- | --- | --- |
| **Breast cancer** |  |  |  |  |  |  |
| 2021 | 14.6 (10.4, 27.1) | 2737 (1950, 5080) | 2354 (1677, 4369) | 383 (273, 711) | 76309 (54366, 141632) | 1853 (1321, 3439) |
| 2026 | 15.8 (10.2, 33.8) | 3035 (1960, 6496) | 2610 (1686, 5587) | 425 (274, 909) | 84615 (54648, 181112) | 2055 (1327, 4398) |
| 2031 | 17.5 (9.9, 42.1) | 3336 (1934, 8223) | 2869 (1663, 7072) | 467 (271, 1151) | 93008 (53918, 229259) | 2260 (1309, 5567) |
| 2036 | 18.5 (9.7, 52.4) | 3638 (1911, 10321) | 3129 (1643, 6772) | 509 (268, 1445) | 101430 (53275, 226733) | 2464 (1294, 6988) |
| **Cervical cancer** |  |  |  |  |  |  |
| 2021 | 10.2 (8.7, 14.4) | 1,912 (1631, 2700) | 1772 (1512, 2503) | 140 (119, 197) | 75080 (64050, 106030) | 1308 (1116, 1848) |
| 2026 | 9.4 (7.8, 13.9) | 1,807 (1499, 2671) | 1675 (1390, 2476) | 132 (109, 195) | 70960 (58870, 104890) | 1236 (1026, 1828) |
| 2031 | 8.7 (7.1, 13.4) | 1,699 (1387, 2617) | 1575 (1286, 2426) | 124 (101, 191) | 66720 (54470, 102770) | 1163 (949, 1791) |
| 2036 | 8.0 (6.3, 12.9) | 1,576 (1241, 2541) | 1461 (1150, 2356) | 115 (91, 185) | 61890 (48730, 99790) | 1078 (849, 1739) |
| **Oral cancer – Male** |  |  |  |  |  |  |
| 2021 | 3.7 (3.0, 4.3) | 702 (569, 816) | 642 (521, 747) | 60 (48, 69) | 27420 (22232, 31881) | 389 (315, 452) |
| 2026 | 3.7 (2.8, 4.5) | 718 (544, 874) | 657 (498, 800) | 61 (46, 74) | 28049 (21254, 34146) | 397 (301, 484) |
| 2031 | 3.7 (2.7, 4.7) | 728 (531, 925) | 666 (486, 846) | 62 (45, 79) | 28438 (20745, 36131) | 403 (294, 512) |
| 2036 | 3.7 (2.5, 4.8) | 732 (494, 949) | 670 (452, 868) | 62 (42, 81) | 28598 (19298, 37069) | 405 (273, 525) |
| **Oral cancer - female** |  |  |  |  |  |  |
| 2021 | 7.2 (6.4, 8.2) | 1358 (1200, 1592) | 1242 (1098, 1457) | 116 (102, 135) | 53049 (46878, 62195) | 752 (664, 881) |
| 2026 | 7.2 (6.1, 8.3) | 1393 (1172, 1595) | 1274 (1072, 1459) | 119 (100, 136) | 54416 (45789, 62304) | 771 (649, 883) |
| 2031 | 7.2 (5.9, 8.3) | 1415 (1152, 1621) | 1295 (1054, 1483) | 120 (98, 138) | 55275 (45002, 63322) | 783 (638, 897) |
| 2036 | 7.2 (5.6, 8.4) | 1427 (1103, 1654) | 1306 (1009, 1513) | 121 (94, 141) | 55744 (43086, 64609) | 790 (610, 915) |
| **Lung cancer - Male** |  |  |  |  |  |  |
| 2021 | 4.5 (3.8, 5.3) | 860 (721, 1006) | 438 (367, 512) | 422 (354, 494) | 25894 (21708, 30288) | 86 (72, 101) |
| 2026 | 4.5 (3.6, 5.4) | 866 (699, 1038) | 441 (356, 528) | 425 (343, 510) | 26075 (21048, 31248) | 87 (65, 109) |
| 2031 | 4.4 (3.3, 5.5) | 865 (649, 1082) | 440 (330, 551) | 425 (319, 531) | 26044 (19536, 32580) | 87 (65, 109) |
| 2036 | 4.3 (3.2, 5.6) | 856 (609, 1107) | 436 (310, 564) | 420 (299, 543) | 25773 (18336, 33336) | 86 (61, 111) |
| **Lung cancer- female** |  |  |  |  |  |  |
| 2021 | 1.99 (1.6, 3.6) | 373 (300, 675) | 190 (153, 344) | 183 (147, 331) | 11231 (9036, 20328) | 37 (30, 68) |
| 2026 | 1.98 (1.6, 3.7) | 381 (308, 711) | 194 (157, 362) | 187 (151, 349) | 11472 (9276, 21408) | 38 (31, 71) |
| 2031 | 1.97 (1.5, 3.8) | 385 (293, 742) | 196 (149, 378) | 189 (144, 364) | 11592 (8820, 22344) | 39 (29, 74) |
| 2036 | 1.96 (1.4, 3.9) | 387 (276, 768) | 197 (141, 391) | 190 (135, 377) | 11652 (8316, 23124) | 39 (28, 77) |

CIR, crude incidence rate per 100000;
Estimated female populations for the years 2021, 2026, 2031, 2036 were 18743719, 19242424, 19543147, and 19744898, respectively; Estimated male populations for the years 2021, 2026, 2031, 2036 were 17718281, 17219576, 16918853, and, 16717102 respectively;
Proportion of breast cancer cases: early stage= 85.98%, late stage= 14.02%; Proportion of cervical cancer cases: early stage= 92.69%, late stage= 7.31%; Proportion of oral cancer cases: early stage= 91.49%, late stage= 8.51%; Proportion of lung cancer cases: early stage= 50.91%, late stage= 49.09%;
A 5-year survival for breast cancer: early stage= 0.7632, late stage= 0.1491 A 5-year survival for cervical cancer; early stage= 0.7318; late stage= 0.0816 A 5-year survival for oral cancer: early stage= 0.6017, late stage= 0.0349; A 5-year survival for lung cancer: early stage= 0.1003, late stage=0.1003
Workload was defined as average number of visits to tertiary centers per case for consultations with oncologists, diagnostic investigations, active treatment and follow-up. Number of visits for Breast Cancer: early stage= 29; late stage= 21; Number of visits for Cervical Cancer: early stage= 40; late stage= 30; Number of visits for Oral cancer: early stage= 40; late stage= 29; Number of visits for Lung Cancer: early stage= 36; late stage= 24;

Table 2 describes the estimated chemotherapy, radiotherapy and surgery requirements for new cases of cancers from the year 2021 to 2036 under different scenarios. The requirement of chemotherapy, radiotherapy and surgery per year will rise by 28% in 2036 compared to 2021 due to increase in new cases of cancer per year. For example, if we assume that 55% of new cancer cases require chemo/radiotherapy and 45% require surgeries, then in the year 2021 for 28785 new cases, 15,832 cases will require chemo/radiotherapy and 12,953 cases will require surgeries. This would translates into 260 chemo cycles/ day, 23 new LINACs, and 1079 surgeries per month. For the same scenario, in the year 2036, to cater the needs of new cancer cases (36776), we will require 332 chemo cycles/ day, 30 new LINACs, and 1379 surgeries per month.

**Table 2.** Estimated chemotherapy/radiotherapy/surgery requirement in Telangana

| Year | New cases | Estimated chemotherapy required |  | Estimated radiotherapy required |  | Estimated cancer surgeries required |  |
| --- | --- | --- | --- | --- | --- | --- | --- |
|  |  | New cases requiring chemotherapy | Chemo cycles/day | New cases requiring RT | LINACs require | New cases requiring surgery | Surgeries/ month |
|  |  | 55% of new cancer cases requiring chemo/radiotherapy* |  |  |  | 45% of new cancer cases requiring surgery* |  |
| 2021 | 28785 | 15,832 (15665, 15999) | 260 (258, 263) | 15,832 (15665, 15999) | 23 (23, 23) | 12,953 (12789, 13120) | 1079 (1066, 1093) |
| 2026 | 31267 | 17197 (17025, 17369) | 283 (280, 286) | 17197 (17025, 17369) | 25 (25, 25) | 14070 (13898, 14242) | 1173 (1158, 1187) |
| 2031 | 33926 | 18659 (18473, 18846) | 307 (304, 310) | 18659 (18473, 18846) | 27 (27, 28) | 15267 (15087, 15447) | 1272 (1257, 1287) |
| 2036 | 36776 | 20227 (20025, 20429) | 332* (329, 336) | 20227 (20025, 20429) | 30* (29, 30) | 16549 (16362, 16737) | 1379* (1363, 1395) |
|  |  | 60% of new cancer cases requiring chemo/radiotherapy |  |  |  | 40% of new cancer cases requiring surgery |  |
| 2021 | 28785 | 17271 (17107, 17435) | 284 (281, 287) | 17271 (17107, 17435) | 25 (25, 26) | 11,514 (11350, 11678) | 960 (946, 973) |
| 2026 | 31267 | 18760 (18588, 18929) | 308 (306, 311) | 18760 (18588, 18929) | 28 (27, 28) | 12507 (12338, 12679) | 1042 (1028, 1057) |
| 2031 | 33926 | 20356 (20179, 20532) | 335 (332, 338) | 20356 (20179, 20532) | 30 (30, 30) | 13570 (13394, 13747) | 1131 (1116, 1146) |
| 2036 | 36776 | 22066 (21882, 22249) | 363 (360, 366) | 22066 (21882, 22249) | 32 (32, 33) | 14710 (14527, 14894) | 1226 (1211, 1241) |
|  |  | 65% of new cancer cases requiring chemo/radiotherapy |  |  |  | 35% of new cancer cases requiring surgery |  |
| 2021 | 28785 | 18710 (18552, 18869) | 308 (305, 310) | 18710 (18552, 18869) | 27 (27, 28) | 10075 (9916, 10236) | 840 (826, 853) |
| 2026 | 31267 | 20324 (20158, 20489) | 334 (331, 337) | 20324 (20158, 20489) | 30 (30, 30) | 10943 (10778, 11109) | 912 (898, 926) |
| 2031 | 33926 | 22052 (21879, 22225) | 362 (360, 365) | 22052 (21879, 22225) | 32 (32, 33) | 11874 (11701, 12047) | 990 (975, 1004) |
| 2036 | 36776 | 23904 (23724, 24085) | 393 (390, 396) | 23904 (23724, 24085) | 35 (35, 35) | 12872 (12691, 13052) | 1073 (1058, 1088) |

LINACs, linear accelerators; RT, radiotherapy;
Chemotherapy requirement calculated based on six cycles per patient assumed; RT services requirement calculated considering 22 RT sessions per patient and 15000 RT sessions per machine assumed;
\*For the year 2036, 55% of new cancer cases (i.e 20227) requiring chemo/radiotherapy and 45% of new cancer cases (i.e. 16549) requiring surgeries would translate into 332 extra chemotherapy cycles per day, 30 new linear accelerators to deliver radiotherapy and 1379 extra surgeries for cancer per month to cater the needs of new cancer case;

Table 3 describes the stakeholders’ perception about the stages of cancer presentation, reasons for delayed presentation, causes of non-compliance, and system level barriers to access cancer care services.

**Table 3.** Stages of cancer presentation, reasons for delayed presentation, causes of non-compliance, and system level barriers: Stakeholders perceptions

| **Major cancers in Telangana and stage at presentation to oncologist (n= 25)** |  |  |  |
| --- | --- | --- | --- |
| Site of Cancer | Stage of Presentation | % of patients | Sector |
| Oral | III | 40 | Public |
|  | II-III | 70-80 | Private |
|  | III | 50 | Private-charitable |
| Breast | III | 30 | Public |
|  | II-III | 70 | Private |
|  | III-IV | 60 | Private-charitable |
| Cervix-uterine | III | 30-40 | Public |
|  | II-III | 80 | Private |
|  | III | 50 | Private-charitable |
| Lung | IV | 40 | Public |
|  | II-IV | 70 | Private |
|  | IV | 90 | Private-charitable |
| **Reasons enumerated by providers for delayed presentation (n= 25)** |  |  | n (%) |
| Lack of awareness of condition & Services |  |  | 21 (84) |
| Stigma associated with diagnosis |  |  | 19 (76) |
| Financial constraints |  |  | 15 (60) |
| Rural background |  |  | 12 (48) |
| Absence of family support/ caregiver to accompany |  |  | 9 (36) |
| Preference to TCAM |  |  | 5 (20) |
| **Causes for non-compliance to recommended therapy or referral advice (n= 25)** |  |  | n (%) |
| Psychological / Attitudinal barriers |  |  | 15 (60) |
| Long distance – travel to higher centers |  |  | 12 (48) |
| Myths and misconceptions |  |  | 4 (16) |
| Accommodation concerns |  |  | 2 (8) |
| **System Level Barriers (n= 25)** |  |  | n (%) |
| Lack of systematic / organized screening |  |  | 15 (60) |
| Effects of pandemic on service delivery |  |  | 13 (52) |
| Inadequate cancer care centers - infrastructure |  |  | 13 (52) |
| Shortage of trained manpower |  |  | 11 (44) |
| Limited supportive care for therapeutic |  |  | 6 (24) |
| Referral pathways - linkages |  |  | 6 (24) |
| Non-availability of essential drugs |  |  | 4 (16) |
| Absence of standardized guidelines / protocols |  |  | 3 (12) |
| Absence of requisite equipment |  |  | 3 (12) |

\*TCAM, traditional, complementary & Alternative medicine;

### Delayed presentation - Lack of awareness and determinants of health seeking behavior

Presentation to a healthcare facility in advanced stages was the common barrier perceived by health care providers at all levels of the health system in both public and private sectors.

84% of the healthcare providers mentioned lack of awareness as the main reason for delayed presentation. Inadequate knowledge on risk factors and screening measures such as procedure for self-examination; negligence of early symptoms and preference of local quacks to allopathic practitioners were the common patient-related barriers encountered by primary healthcare providers. Inadequate knowledge about the condition and available health care services was evident across patient-groups, with women and those from rural backgrounds being disproportionately affected.

*“We did not know earlier, approached several centers over the period of one year, used herbal medicines, have also been to Shivamogga in Karnataka” – [Patient: 17, Stomach Cancer]*

*I never knew about cancer, until I was diagnosed. I thought lump underneath breast was normal for pregnant and lactating women and believed cancer was a result of straining my voice at workplace – [Patient: 1, Breast cancer]*

Educational status was associated with awareness on cancers, wherein none of the patients who were educated up to the senior secondary was aware of cancer as against 83.3% of patients among those who were graduates or more educated. The median duration from the initial identification of symptoms to receiving diagnosis was greater amongst patients with lower educational attainment and those with less awareness on cancer compared to those counterparts with higher educational status and subsequently, greater awareness.

Medical officers both at primary and secondary levels of the state’ s health system were concerned that social determinants such as literacy status, gender, religion, economic status and location of regular habitation (rural/urban/tribal) have a synergistic effect on individual health seeking behaviour in general, and cancer prevention, in particular. The association of socioeconomic status with age at marriage, menstrual hygiene and sanitation and in turn, their role in infection-related cancers were enumerated by physicians and nurses at the secondary level of the public health system.

### Delayed presentation - Stigma and misconception

Stakeholders at the community level emphasized that stigma was the prime reason responsible for the late presentation of cancer patients at health centers as villagers have a taboo and apprehension that revealing their diagnosis may lead to them being ‘ labelled’ by the community which results in them facing hardships due to societal norms.

### *Delayed presentation -* Financial barriers

While all the providers at tertiary level perceived financial constraints as a cause for delayed presentation, only 28.6% primary healthcare personnel had this perception. Also, 90% providers in the older age cohort perceived financial barriers delay healthcare-seeking as against 40% providers in the younger age cohort. Although diagnostic investigations and treatment would be provided without any user-fee in the public health facilities, 60% stakeholders contended that indirect costs in terms of loss of wages and productive work hours is a constraint for patients from families below the poverty line, more so if patient or primary caregiver is the sole earning member. Moreover, direct non-medical costs for transportation and accommodation ought to be borne by the patient or his family and affordability is a fundamental challenge for those from peripheral areas, as cancer treatment lasts for longer duration.

While 60% patients who had knowledge on cancer were not covered under any insurance scheme, 66.7% of patients who were not aware of the condition had govt. sponsored insurance scheme and these differences were statistically significant. 47.1% of the 17 cancer patients interviewed reported receiving treatment under the state-sponsored insurance scheme. As elucidated by providers, direct non-medical costs such as expenses for travel and accommodation were the major concerns of patients and their families. Owing to the unanticipated diagnosis, patients resorted to distress financing for funding current treatment. Out-of-pocket expenditure varied from 40% to 70%, due to limited coverage of cancer-related therapies under both state-sponsored and private medical insurance schemes.

### Delayed presentation - Place of habitation and absence of support

While 85.7% of seven tertiary level healthcare providers opined that rural background is a determinant of delayed presentation, only 40% and 14.3% of providers at the secondary and primary levels respectively, had the same concern. All the private sector healthcare practitioners perceived rural background as a barrier for early presentation compared to 35% of those serving in the public sector. Caregivers from rural backgrounds despite poor literacy had extreme difficulty navigating patients to the tertiary cancer centers aggregated in urban areas. Loss of employment and livelihood for the entire duration of treatment was reiterated by caregivers. Caregiving resulted in role-conflicts and primary caregivers also experienced emotional trauma as some of them did not reveal the exact diagnosis to either the patient or other members of the family.

### Non-compliance to recommended therapy

Oncologists mentioned that patients’ perception of cancer treatments and subsequent side-effects such as loss of hair during chemotherapy, propel them to default the initiated therapy and search for alternatives under the assumption that the recommended treatment is ineffective. Perceived barriers behind treatment defaulting include beliefs that cancer is incurable, or radiotherapy would result in burns or death of the patient and that cancer patients receiving treatment should consume only plant-based diet.

*“Once patients return home and after the first cycle, they feel they are completely cured and don’ t return for the next cycle on time, until the disease progresses and return when they start experiencing discomfort/difficulty*.*”* – Provider 3

“Why did this come to me? No one in our family has this problem. Then why did this happen to me?” - [Patient: 5, Breast Cancer]

60% healthcare providers expressed psychological concerns such as anxiety as a cause for non-compliance. Social circumstances such as absence of family support and /or caregiver to accompany the patient and distant location of diagnostic and treatment centers were considered by providers as the causes for non-compliance to referral advice and recommended therapy. While 20% of oncologists perceived supportive care as a barrier, 66.7% of nurses along with both the palliative care physicians and the stakeholder from community-based NGO perceived it as a barrier for patients.

While cancer patients who experienced accommodation/shelter barriers had an average duration of 4.5 months from noticing symptoms to being diagnosed with cancer, this duration was only 1.5 months for those without such concerns. In comparison to patients without transportation barriers, those with such challenges had a longer interval between first identification of symptoms and receiving the confirmatory diagnosis.

### System-level barriers

System-induced barriers mainly consisted of issues concerning infrastructure and workforce essential for service-delivery (including screening), in addition to ill-effects of the pandemic. Lack of systematic/ organized screening programs, effect of the COVID pandemic and inadequate infrastructure were the three most important barriers identified by health care providers (Table 3). Health care providers at the primary level identified challenges in the implementation of the National Programme for prevention & Control of Cancer, Diabetes, Cardiovascular Diseases & stroke (NPCDCS) program which impacts cancer care services in Telangana: (i) ANMs in few districts not trained in screening for cervical cancer; (ii) Risk of missing cases at the primary level; (iii) Complex reporting formats; (iv) Multiple roles assigned to ANMs under the existing health care programmes (reproductive health programs and NPCDCS) in the scheduled work hours and (iv) Reduced workforce efficiency. At the primary level, screening activities have had been initiated in the state but ANMs only in certain districts were trained in early detection of all the three common cancers – breast, cervical and oral. Although, ANMs were conducting oral visual examination and clinical breast examination, training in VIA-based (Visual Inspection with Acetic acid) screening for early detection of cervical cancer hadn’ t been completed in some of the districts.

Absence of histopathology labs, diagnostic equipment and chemo drugs for onco-specific treatment at the district level, including medical colleges in the public sector was identified as the principal barrier by providers at the secondary level. At the tertiary level, absence of designated areas for mixing of chemo-drugs and deficit of protective gear required during the process were some of the challenges faced by nursing personnel in the public sector. Despite the presence of shift-duties, due to extensive patient-volumes at the tertiary cancer centers in the public sector, provision of quality care was a perceived barrier. In addition to patient-care, administration of chemotherapy and diet-maintenance, nurses were engaged in documentation and ‘ clerical work’, exacerbating the problem.

*Patient mainly expect us to listen to their problems. But we can’ t spend enough time with each of them*.*” – [Provider: 12]*

### System level barriers - Patient care pathways

Absence or ill-defined standardized pathways for cancer management (aimed to ensure timely diagnosis followed by prompt and appropriate treatment) was another important barrier highlighted by healthcare providers. Physicians at the primary healthcare level opined that referring to secondary level health centers is of limited use as there is a shortage of specialists and equipment for confirmatory diagnosis and staging. At the secondary level, medical officers and civil assistant surgeons highlighted the need for standard protocols for providers to ensure appropriate referral of screen-positives and suspected cases.

### Major cancers in Telangana and stage at presentation to oncologist (n= 25)

The required number of human resources and equipment for new cancer cases from year 2021 to 2036 are summarized in Fig 1. There is a steady increase in the requirement of human resources and equipment per year with rising number of new cancer cases. In the year 2036 (compared to the year 2021), there will be 28% increase in the requirement of human resources (such as Pathologist, Surgical oncologists, Medical oncologists, Palliative care specialists, Clinical pharmacists, Medical physicists, Radiation / Clinical oncologist, and Radiotherapy technicians) and equipment (Megavoltage teletherapy unit, Brachytherapy unit, CT simulator) per year from 2021 to 2036.

**Fig 1.**
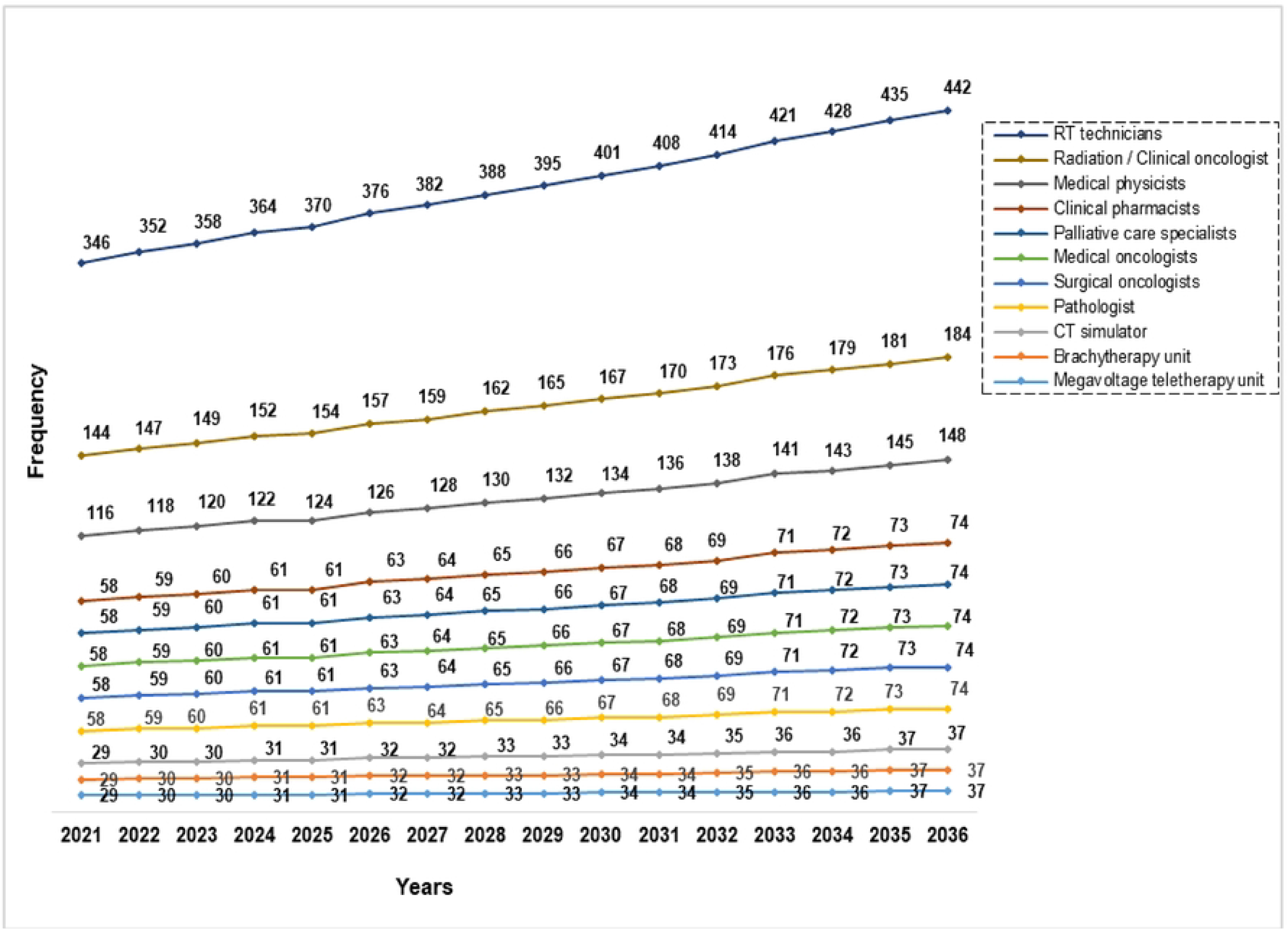
Human Resources and Equipment requirement for new cancer cases from 2021-2036. RT, radiotherapy; CT, computed tomography; Estimated new cases for the year 2021, 2022, 2023, 2024, 2025, 2026, 2027, 2028, 2029, 2030, 2031, 2032, 2033, 2034, 2035, and 2036 are 28785, 29268, 29757, 30253, 30757, 31267, 31784, 32308, 32840, 33379, 33926, 34480, 35042, 35612, 36190, and 36776, respectively. The recommended norms per 1000 new cancer cases[12] for Megavoltage teletherapy unit, Brachytherapy unit, CT simulator, Pathologist, Surgical oncologists, Medical oncologists, Palliative care specialists, Clinical pharmacists, Medical physicists, Radiation / Clinical oncologist, and Radiotherapy technicians are 1, 1, 1, 2, 2, 2, 2, 2, 4, 5, and 12, respectively.

Experts reported leakage of patients from screened positive till treatment completion in Fig 2. Major leakage of patient was highlighted between ‘ screened-positive’ to ‘ diagnosis confirmation’ step. Overall from ‘ screened-positive’ till ‘ treatment completion’, patient leakage varied from ∼70% to 90%. The findings were relatively same when we developed patient leakage funnels separately for cervical and breast cancers (S1A and B Fig).

**Fig 2.**
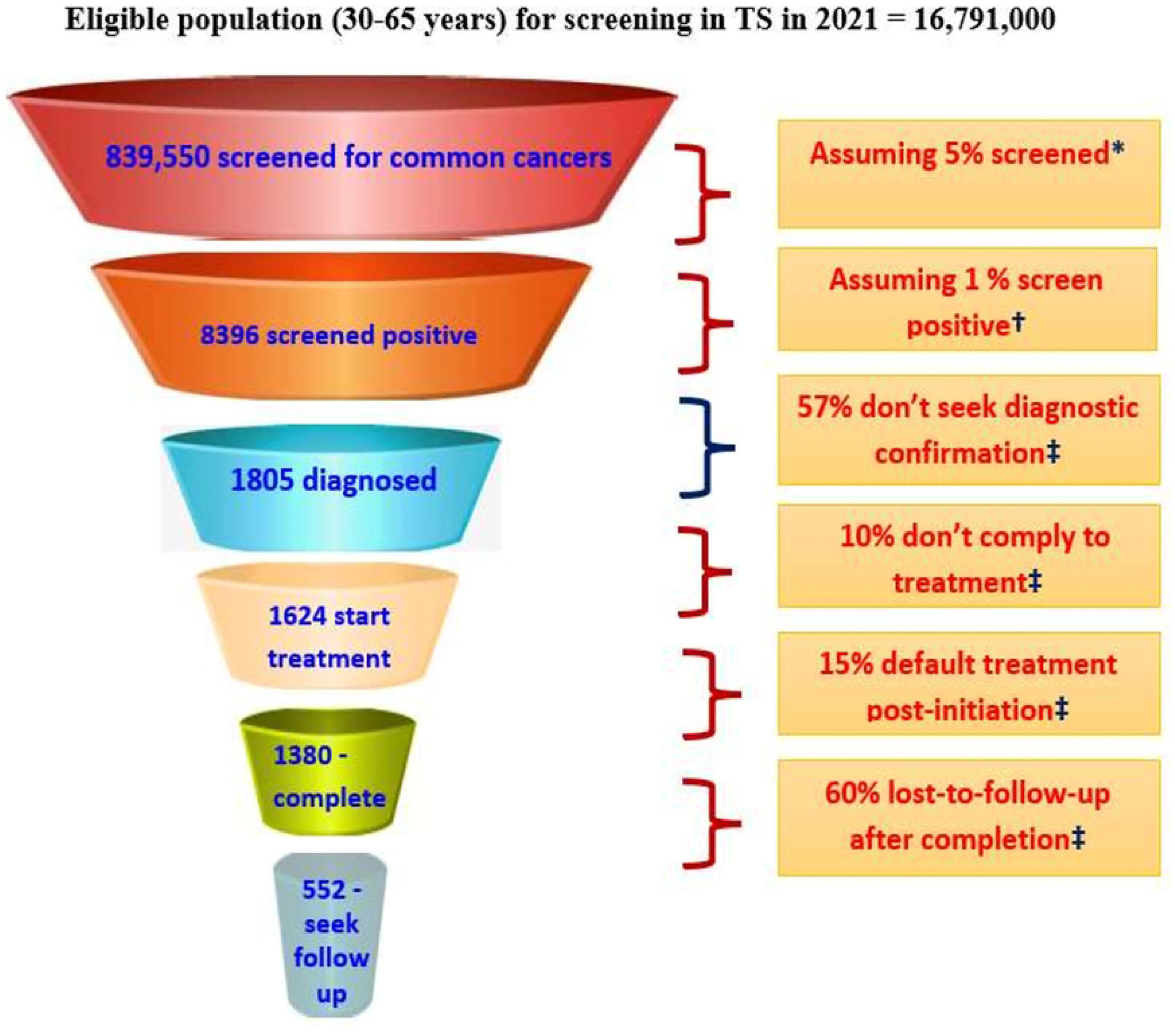
Patient leakage at various stages in the continuum of cancer care for breast, cervical and oral cancer together. *Extrapolated from percentage screened amongst 30-49 years age-group according to NFHS-5 †Based on age-truncated case detection rate of cancers (of all sites) in India [13] ‡3610 (43%) seek diagnostic confirmation, assuming 50% of these to be cases. ‡Based on interviews with oncologists. (a) Total patient loss (from screened positive till treatment completion) = [8396-552]/[8396]= 93.4%; (b) Assuming all screened positive seek diagnostic confirmation & 50% of which test positive, patient loss= [(8396/2)-552] / [8396/2] = 87.0%; (c) From confirmed diagnosis till treatment completion, patient loss =(1805 – 552) /1805= 69.4%

## Discussion

We conducted a situational analysis of cancer care activities for the state of Telangana to identify potential public health value additions and complement the cancer care initiatives of the State Government. We found multiple challenges in cancer care activities that need attention to influence cancer prevention in TS. These are: (i) ∼28% increase in the number of new cancer cases and 244% increase in the number people living with cancer from 2021 to 2036; (ii) Proportionate increase (i.e. 28%) in the workload on the healthcare system (for example number of patients’ visits, chemotherapy sessions /radiotherapy sessions, number of surgeries per day) from 2021 to 2036; (iii) 28% higher annual requirement of human and equipment from 2021 to 2036 to cater the increasing workload; (iv) Suboptimal treatment seeking behavior among patients (delayed presentation to hospital for cancer care) accompanied with non-compliance to treatment due to individual as well as system level barriers; (v) Major patient leakage of 70 to 90% from screening till treatment completion for common cancers. To our knowledge, this is the first study from India which using mixed method has assessed human and equipment requirement for the rising burden of cancer and highlighted the individual- and system-level barriers for major patient leakage across cancer care continuum.

### Delayed Access to Healthcare system

This study has identified several public health additions that should be addressed to further catalyze the prevention and control of cancers in Telangana. Stakeholders’ repeatedly observed the delayed presentation of cancer patients which is the main reasons for the poor survival of cancer patients in India compared to their European counterparts. Most of the stakeholders reported lack of awareness or misconception about cancer, its risk factors followed by financial constraints. This finding is consistent with the results reported from Orissa (one of the neighboring state of Telangana) where researchers quantified the delays in care-seeking for signs and symptoms related to cancer [14]. They found that the first step in the pathway-to-care was sharing symptoms or signs with family members and friends, and on average, took 271 days before steps toward diagnosis were taken [14]. Lack of knowledge, fear, and stigma related to cancer were highlighted as the key factors influencing this delay [14]. The cancer literacy plays a key role in cancer outcomes and the government should consider investing significantly in cancer prevention education. Our findings also highlight: (i) To identify effective measures to promote cancer literacy across all stakeholders; (ii) To monitor and evaluate these strategies to achieve optimum benefit; (iii) In-depth understanding of socio-cultural factor such as myths, misconception etc. and other barriers to access cancer care services.

Stakeholders also mentioned about the financial constraint as one of the major deterrents for early access to healthcare facility; and therefore cancer prevention efforts should ensure access to affordable treatment. Cancer patients in India incur heavy out-of-pocket expenditures [15, 16]. Several central as well as state insurance schemes exist for cancer patients in Telangana to cover different aspects of cancer care. Some of the schemes are: Rashtriya Aarogya Nidhi Health Minister’ s Cancer Patient Fund, The Employees’ State Insurance scheme, Telangana state Aarogyasri scheme, The Employees and Journalists Health Scheme of Telangana etc. that cover cancer care including specialized investigation as well as surgery, chemotherapy, radiotherapy, and palliative chemotherapy [17]. Government should facilitate the regular monitoring and evaluation of these schemes to ensure that these schemes cover all the components across cancer care continuum. Data from National Sample Survey-75th round points to 70.3% of rural and 37.3% urban households in Telangana being covered under a government sponsored health insurance scheme. In case of hospitalized cancer patients, 83% were admitted at private hospitals from those hailing from rural Telangana as against 70% from those belonging to urban areas. Overall, 79% of hospitalized cancer patients in Telangana were admitted in private hospitals who might have incurred huge out of pocket expenses if had not been covered under any insurance scheme [18].

### Patient leakage across cancer care continuum

This study also highlighted the variable leakage of patients across multiple stages in cancer care continuum with major leakage happening from screening to diagnosis confirmation. The loss of patients across cancer care continuum could be explained by factors operating at three levels: a) Reduced generation and dissemination of scientific knowledge regarding interventions, practices and services; b) Inadequately prepared healthcare system delivering inefficient services; and c) Poor demand and uptake of cancer services due to social, cultural and economic factors. Rigorous research across these three factors could identify needs and gaps in the existing cancer care system, which if addressed, could catalyze the cancer care services in the state of Telangana. As suggested previously[19], WHO’ s six building blocks of the health system framework consisting of information, medical products and technologies, service delivery, health workforce, financing, and leadership and governance can be used to identify research needs to promote generation and dissemination of scientific knowledge regarding interventions, practices and services and strengthen healthcare system to deliver inefficient services. Additionally, poor demand and uptake of cancer services due to social, cultural and economic factors can be mitigated by tackling context-specific culturally relevant cancer prevention education.

The government of Telangana has been showing a strong commitment and taking major initiatives to strengthen the cancer care services across the cancer care continuum (S4 table). Telangana government is attempting to develop a decentralized cancer care system comprising stakeholders from public, public-private partnership, and non-governmental institutes to provide equitable, accessible, affordable, safe and effective cancer care services [20]. Government is planning to have an apex center in Hyderabad (Level-1 cancer care center with facilities for advanced diagnostics, treatment, research, and education) in addition to existing MNJ Cancer Hospital, Nizam’ s Institute of Medical sciences, and several private hospitals in Hyderabad.

Medical colleges at Adilabad, Nizamabad, Mahbubnagar and Warangal are being strengthened as Level-2 centres to provide diagnostic and treatment services in all modalities such as surgical care, radiotherapy, and chemotherapy. Diagnostic hubs at the level of district (Level-3 facilities) to provide definitive diagnostics, day-care and palliative therapy. Government is actively promoting the training of ANMs at sub-center (SC) and medical officer at primary health center (PHC), and community health center (CHC), to efficiently utilize the extensive network of SCs, health wellness canters (HWCs), PHCs, and CHCs to create awareness and provide basic screening services to ensure early diagnosis and prevent future cases. Additionally, several initiatives (by governmental and non-governmental organizations) are being carried out to -raise cancer literacy, -address fear and stigma, -formulate, -disseminate, and -implement evidence-based guidelines for screening, diagnosis and treatment (including palliative care), -increase cancer care centers/institutes and -address shortages in healthcare facilities, medical resources, and healthcare professionals to ensure timely care-seeking and follow-up (S4 Table). These newer strategies should be accompanied with rigours monitoring and evaluation tools to identify appropriate service delivery and financing models.

### Strengths

For in-depth interviews, we have considered cancer patients (with different organ involvement and were at different stages) and healthcare providers working at different levels of healthcare to understand the complete dimension of cancer care services. This activity also helped us to develop patient leakage funnel in the absence of scientific evidence to appreciate at what levels of care patient leakages occur due to sub-optimal efforts and therefore required extra efforts.

### Limitations

The findings highlighted in this paper should be considered in the light of limitations. First, the number of patients as well as healthcare providers interviewed were very small. However, the in-depth interviews of these stakeholders helped us to highlight the issues in cancer care (at patient-as well as system-level) which need urgent attention to strengthen the cancer care services; Second, we could not find out the existing number of human resources and equipment in the state of Telangana due to limited access to public as well private healthcare institutes; therefore could not calculate the exact current deficit of these resources. However, interviews with different stakeholders working at different level of healthcare gave us the glimpse of limited availability of these resources across various sectors. Third, we were only able to conduct phone interviews due to the COVID-19 pandemic, which may have led to altered responses. However, the interviewers were adequately trained before the interviews to extract relevant information from the stakeholders. Fourth, we made several assumptions and used anecdotal evidence to calculate the expected burden for cancer in next 15 years as well as to highlight patient leakages in cancer care funnel. Although we were unable to calculate the exact numbers for projections as well as patient leakages, we have delineated dual nature of problem in cancer care: rising number of cases accompanied by poor availability and/or utilization of healthcare system.

### Public health implications

Amidst globalization, many lower middle income countries (including India) are experiencing urbanization, dietary transition (from fruits and green leafy vegetables to calorie-rich and nutrient sparse fried food), and changes in environmental exposures which could increase the burden of overweight, obesity, and non-communicable diseases including cancer. Policymakers in India are struggling to identify optimal strategies to allocate limited resources for non-communicable diseases as these are already being utilized for communicable diseases and maternal and child health. This study very clearly highlighted the dual problems for cancer care: rising burden and delayed access to healthcare system, in a population which is experiencing epidemiological transition (aging and lifestyle changes). Additionally, we have highlighted the specific areas in the cancer care continuum which needs attention (for example more implementation research) to facilitate early diagnosis of cancer and thereby better prognosis. Improving the health of transitional population, for instance through cancer prevention and control will positively impact economic productivity in India. This will be particularly significant as cancer develop at earlier ages in south Asians (during their economic productive years).

## Conclusion

Telangana state has made commendable efforts towards improving the health profile of its citizens. However, there is ample scope for improvement in the delivery of comprehensive cancer care services in the state. This study has highlighted multiple challenges in cancer care services: Rising burden of cancer cases; increasing requirement of human and equipment; Delayed access; Geographical disparity; Major patient leakage across cancer care continuum. Implementation research focusing on these challenges/barriers is required to identify the optimum strategy to prevent cancer and promote the uptake of cancer care services.

## Data Availability

Data cannot be shared publicly because of Institutional Data Privacy Policies. Data are available from the Indian Institute of Public Health, Hyderabad Institutional Data Access / Ethics Committee (contact via) for researchers who meet the criteria for access to confidential data.

## Acknowledgements

The authors have no acknowledgments to declare.

## Authors’ contribution

HM and GVS formulated the research question. All authors contributed to the design of the study. HM, NR, NGM, and URP prepared the protocol. GVS, MJ, ST and URP supervised the study. HM, NR and NGM curated the data. HM, NR, and NGM conducted the formal data analysis. HM wrote the first draft of the paper. All authors edited the subsequent drafts and approved the final version of the paper.

## Supporting information

**S1 Table. Estimated magnitude of cancers in Telangana for next 15 years**

**S2 Table. Socio-demographic and designation of healthcare providers**

**S3 Table. Socio-demographic profile of patients (n= 17)**

**S4 Table: Cancer care initiatives in the state of Telangana**

**S1A Figure. Patient leakage at various stages in the continuum of breast cancer care**

**S1B Figure. Patient Leakage at various stages in the continuum of cervical cancer care**

**S1 Appendix. The Telangana Cancer Control Study Group**

## References

[1] Dandona L, Dandona R, Kumar GA, Shukla DK, Paul VK, Balakrishnan K, et al. Nations within a nation: Variations in epidemiological transition across the states of india, 1990-2016 in the global burden of disease study. The Lancet. 2017;390:2437–60. doi:10.1016/S0140-6736(17)32804-0

[2] The burden of cancers and their variations across the states of india: The global burden of disease study 1990-2016. Lancet Oncology. 2018;19:1289–306. doi:10.1016/s1470-2045(18)30447-9

[3] National centre for disease informatics and research, bengaluru. Report of national cancer registry programme (icmr-ncdir), bengaluru, india 2020.

[4] Sarin R, Khandrika L, Hanitha R, Avula A, Batra M, Kaul S, et al. Epidemiological and survival analysis of triple-negative breast cancer cases in a retrospective multicenter study. Indian Journal of Cancer 2016;53:353–9. doi:10.4103/0019-509x.200682

[5] Chalkidou K, Marquez P, Dhillon PK, Teerawattananon Y, Anothaisintawee T, Gadelha CA, et al. Evidence-informed frameworks for cost-effective cancer care and prevention in low, middle, and high-income countries. Lancet Oncology. 2014;15:e119–31. doi:10.1016/s1470-2045(13)70547-3

[6] Sivaram S, Majumdar G, Perin D, Nessa A, Broeders M, Lynge E, et al. Population-based cancer screening programmes in low-income and middle-income countries: Regional consultation of the international cancer screening network in india. Lancet Oncology. 2018;19:e113–e22. doi:10.1016/s1470-2045(18)30003-2

[7] Centers for disease control and prevention. Gats 2 global adult tobacco survey gats objectives gats 2 highlights, 2016. [accessed on: 2021 mar 14].

[8] Telangana state portal state-profile. [accessed on: 2021 sep 3].

[9] Icmr-national center for disease informatics and research. Profile of cancer and related factors - telangana. 2020

[10] Population projections for india and states 2011-2036. New delhi; [accessed on: 2021 aug 28].

[11] Sankaranarayanan R, Swaminathan R, Brenner H, Chen K, Chia KS, Chen JG, et al. Cancer survival in africa, asia, and central america: A population-based study. Lancet Oncol. 2010;11:165–73. doi:10.1016/s1470-2045(09)70335-3

[12] Daphtary M, Agrawal S, Vikram B. Human resources for cancer control in uttar pradesh, india: A case study for low and middle income countries. Frontiers in Oncology. 20144. doi:10.3389/fonc.2014.00237

[13] Shridhar K, Dey S, Bhan CM, Bumb D, Govil J, Dhillon PK. Cancer detection rates in a population-based, opportunistic screening model, new delhi, india. Asian Pacific Journal of Cancer Prevention. 2015;16:1953–8. doi:10.7314/apjcp.2015.16.5.1953

[14] D W, S P. Patient pathways to cancer diagnosis and delay in care seeking: An exploratory study in odisha (india).. European Journal of Cancer Care. 2014;23:1–23. doi:10.1111/ecc.12226

[15] Pramesh C, Badwe RA, Borthakur BB, Chandra M, Raj EH, Kannan T, et al. Delivery of affordable and equitable cancer care in india. The Lancet Oncology. 2014;15:e223–e33

[16] Mahal A, Karan A, Fan VY, Engelgau M. The economic burden of cancers on indian households. PloS ONE. 2013;8:e71853

[17] The annual statistical report 2017-18 government of telangana aarogyasri health care scheme (quality medicare for all). [accessed on: 2021 aug 28].

[18] Ministry of statistics and programme implementation national statistical office. Key indicators of social consumption in india: Health nss 75th round july 2017-june 2018..

[19] Krishnan S, Sivaram S, Anderson BO, Basu P, Belinson JL, Bhatla N, et al. Using implementation science to advance cancer prevention in india. Asian Pacific journal of cancer prevention 2015;16:3639–44. doi:10.7314/apjcp.2015.16.9.3639

[20] Government of telangana and tata trusts signed mou to provide state-wide cancer care: Press releases; [accessed on: 2021 may 11].

